# SEETrials: Leveraging Large Language Models for Safety and Efficacy Extraction in Oncology Clinical Trials

**DOI:** 10.1101/2024.01.18.24301502

**Authors:** Kyeryoung Lee, Hunki Paek, Liang-Chin Huang, C Beau Hilton, Surabhi Datta, Josh Higashi, Nneka Ofoegbu, Jingqi Wang, Samuel M Rubinstein, Andrew J Cowan, Mary Kwok, Jeremy L. Warner, Hua Xu, Xiaoyan Wang

## Abstract

**Background:** Initial insights into oncology clinical trial outcomes are often gleaned manually from conference abstracts. We aimed to develop an automated system to extract safety and efficacy information from study abstracts with high precision and fine granularity, transforming them into computable data for timely clinical decision-making.

**Methods:** We collected clinical trial abstracts from key conferences and PubMed (2012-2023). The SEETrials system was developed with four modules: preprocessing, prompt modeling, knowledge ingestion and postprocessing. We evaluated the system’s performance qualitatively and quantitatively and assessed its generalizability across different cancer types— multiple myeloma (MM), breast, lung, lymphoma, and leukemia. Furthermore, the efficacy and safety of innovative therapies, including CAR-T, bispecific antibodies, and antibody-drug conjugates (ADC), in MM were analyzed across a large scale of clinical trial studies.

**Results:** SEETrials achieved high precision (0.958), recall (sensitivity) (0.944), and F1 score (0.951) across 70 data elements present in the MM trial studies Generalizability tests on four additional cancers yielded precision, recall, and F1 scores within the 0.966-0.986 range. Variation in the distribution of safety and efficacy-related entities was observed across diverse therapies, with certain adverse events more common in specific treatments. Comparative performance analysis using overall response rate (ORR) and complete response (CR) highlighted differences among therapies: CAR-T (ORR: 88%, 95% CI: 84-92%; CR: 95%, 95% CI: 53-66%), bispecific antibodies (ORR: 64%, 95% CI: 55-73%; CR: 27%, 95% CI: 16-37%), and ADC (ORR: 51%, 95% CI: 37-65%; CR: 26%, 95% CI: 1-51%). Notable study heterogeneity was identified (>75% *I*^2^ heterogeneity index scores) across several outcome entities analyzed within therapy subgroups.

**Conclusion:** SEETrials demonstrated highly accurate data extraction and versatility across different therapeutics and various cancer domains. Its automated processing of large datasets facilitates nuanced data comparisons, promoting the swift and effective dissemination of clinical insights.

## INTRODUCTION

Multiple myeloma (MM) remains an incurable malignancy, marked by elevated rates of relapse and therapy resistance despite significant advancements in treatment options and survival rates over the last 15 years ^1^. Furthermore, the introduction of numerous new therapies has led to uncertainty in therapy selection and sequencing, compounded by limited consensus guidelines and recent systematic literature reviews ^2, 3^. Effective clinical decision-making is supported by a robust evidence base, typically sourced from clinical trials ^4^. The prompt dissemination of clinical trial findings, including both risks and benefits, is fundamental for informed patient care ^5^ and crucial in refining patient recruitment strategies for subsequent trial phases—particularly in urgent medical fields like oncology ^6^.

Conference abstracts often serve as crucial sources of initial findings, unveiling clinical trial results and providing insights into the efficacy and safety of investigational treatments. Yet, the initial disclosures via conference abstracts present retrieval and integration challenges, with over half of study outcomes and nearly a third of randomized trial results from these abstracts not advancing to full publication ^7–9^. In addition, publication bias disproportionately excludes "not positive, positive but not statistically significant, or negative" results ^10, 11^ despite their critical influence on clinical decision-making. Recognizing these issues, extracting data from both journal articles and conference abstracts is essential for thorough evidence synthesis and for facilitating robust comparisons. Nevertheless, the extraction of clinical outcomes for large-scale analysis can be formidable when relying solely on manual methods.

Advancements in Natural Language Processing (NLP) have notably enhanced medical research through the extraction of data from unstructured texts ^12–14^. Generative Pre-trained Transformer (GPT) models, a subset of large language models (LLMs), have shown exceptional proficiency in contextual understanding and textual data processing ^15–18^. Various GPT models have been examined for information extraction from clinical documents ^16, 19–21^. In the clinical trial domain, LLMs show promise in trial information retrieval ^22^, criteria text generation ^23^, and clinical trial eligibility criteria analysis ^24^. While LLMs have been utilized to support systematic reviews ^25^, the application of LLMs to extract clinical trial outcomes from conference abstracts, especially in the MM domain, remains largely unexplored.

In this study, we present "SEETrials," a cutting-edge LLM-based pipeline specifically designed for the automatic extraction of safety and efficacy data from a broad spectrum of conference and journal abstracts. By focusing on MM clinical trials and concentrating three recent classes of interventions including chimeric antigen receptor T cell (CAR-T) therapy, bispecific antibodies (BsAbs) therapy, and antibody-drug conjugates (ADC) studies, we demonstrate the practical utility of our approach in enhancing clinical decision-making. Our system also has been shown for its applicability across various cancer trial studies.

## MATERIALS AND METHODS

### Data

For the model development, evaluation, and data analysis, we collected a total of 245 MM clinical trial abstracts (2012-2023) from various drug groups, focusing on five different therapies. The sources included the American Society of Clinical Oncology (ASCO: https://ascopubs.org/jco/meeting ), American Society of Hematology conferences (ASH: https://ashpublications.org/blood/issue/142/Supplement%201 ), and PubMed (https://pubmed.ncbi.nlm.nih.gov/ ) websites. The selection was based on keywords (“clinical trial” AND “each therapy”).

The therapies covered in this study are 1) chimeric antigen receptor T cell (CAR-T) therapy 2) Bispecific Antibodies (BsAbs) therapy 3) Antibody-drug conjugates (ADC) therapy 4) Others including Cereblon E3 ligase modulator therapy (CELMoD), Histone deacetylase inhibitor (HDACi) and immune checkpoint inhibitors (ICI). The breakdown of abstract numbers for each therapy and phase is presented in Supplement Table 1.

Of these, 93 abstracts concerning CAR-T, BsAbs, and ADC therapies were earmarked for in-depth quantitative comparison analysis. Exclusion criteria included studies without reported outcomes or those superseding initial results with expanded trial cohorts. A manual review was conducted to validate the extracted data for quantitative analysis.

An additional 115 abstracts across four other cancers—breast, lung, lymphoma, and leukemia— were collected to assess the system’s generalizability. This included acute lymphocytic leukemia (7), acute myelocytic leukemia (6), chronic lymphocytic leukemia (7), and chronic myeloid leukemia (5).

### SEETrials

We developed SEETrials using GPT-4 to extract clinical trial details from annual conference presentations and published journal abstracts. The system’s architecture is delineated in Figure 1, with each step detailed in Supplement Methods. In summary, SEETrials is comprised of four modules.

1) Pre-processing: This initial module is responsible for the systematic collection of abstracts from the ASCO and ASH conferences, as well as the PubMed database. Within this stage, any tables present in the conference abstracts are meticulously converted to a text format to facilitate further analysis.
2) Knowledge ingestion: We integrated oncology clinical trials’ background knowledge such as the definition and examples of safety and efficacy outcomes into the prompt to enhance the LLM’s analytical capabilities.
3) Prompt modeling: The third module focuses on creating tailored prompts that enable the precise extraction and consolidation of data. These prompts are designed to guide the LLM in identifying and merging relevant trial outcomes effectively.
4) Post-processing: In the final module, the extracted outcome data are refined and structured to suit the requirements of subsequent data analysis tasks.

**Figure 1.**
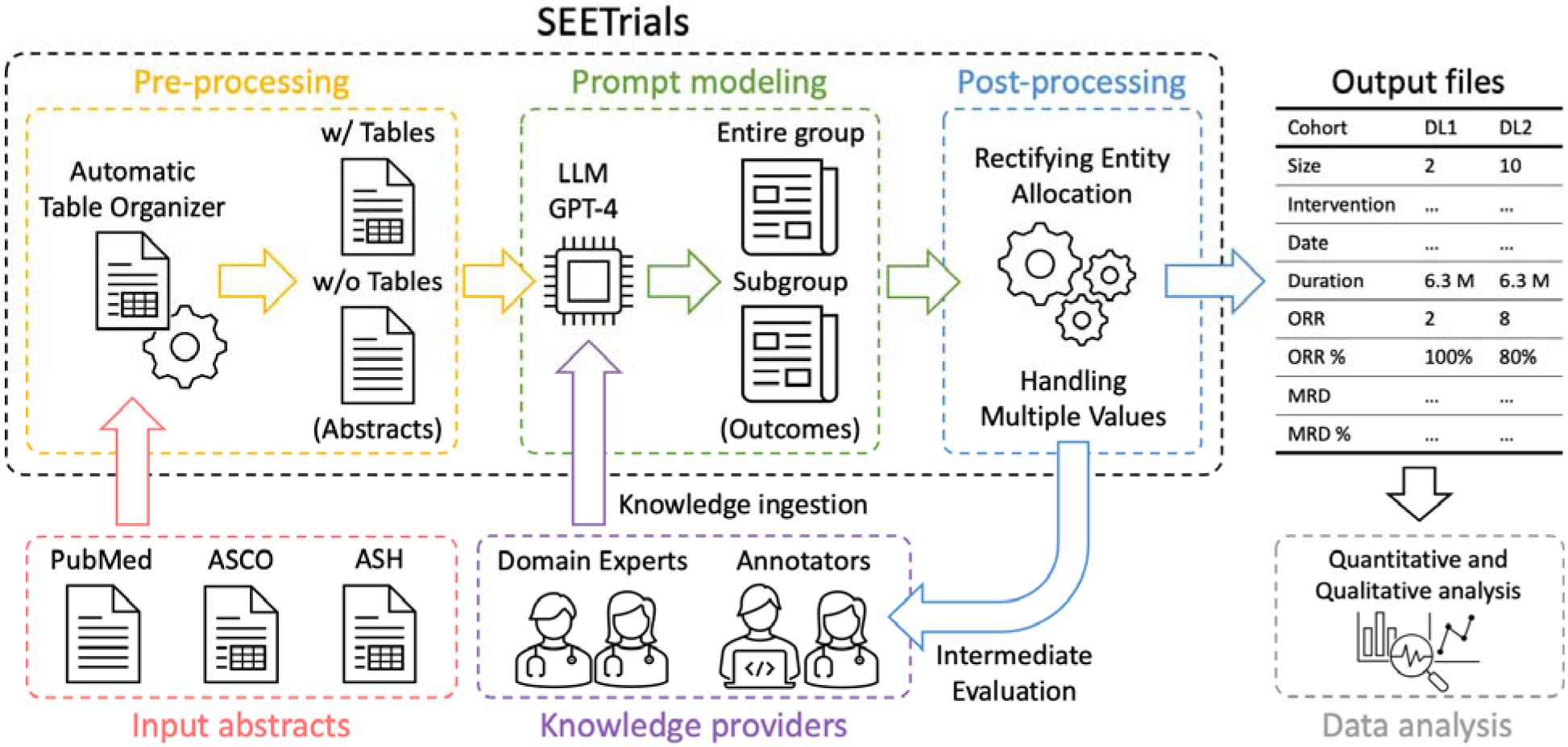
SEETrials system overview. SEETrials is an autonomous system designed to extract critical details from clinical trial studies presented at annual conferences and published journal abstracts. Utilizing the capabilities of GPT-4, the system is delineated in subsequent sections, with a schematic overview of its components. ASCO, American Society of Clinical Oncology; ASH, American Society of Hematology; LLM, Large language model; GPT, Generative Pre-trained Transformer; ORR, overall response rate; MRD, minimum residual disease.

### Evaluation

The evaluation took place using two approaches: Quantitative and qualitative evaluation. For Quantitative evaluation, precision, recall, and F1 scores of SEETrials are reported for each abstract. For qualitative evaluation, predominantly appearing error types were evaluated.

### Data Analysis

In the descriptive data analysis, we extensively scrutinized the distribution of entities and provided a quantitative overview, encompassing both the absolute number and the percentage of abstracts where each entity is mentioned out of the total abstracts analyzed for each therapy. This analysis was performed in conjunction with the categorization of entities based on different trial phases.

We also conducted the quantitative statistical analysis using R software (version 4.0.0), employing the meta, metasens, and metafor packages. A random-effects model was employed, weighting studies by the inverse variance method to favor large population studies. Logit transformations normalized the proportions of overall response rates (ORR), complete response (CR), Neutropenia (≥Grade 3), and Cytokine Release Syndrome (CRS) for effect size pooling. Subgroup analysis was performed for each therapy to investigate treatment effects comprehensively. Statistical heterogeneity among studies was assessed using the *I^2^* inconsistency index and Cochran Q test. Effect sizes were reported with 95% confidence to address variability. Two-sided P values with a significance level of P < 0.05 determined statistical relevance.

## RESULTS

### Evaluation of the SEETrials

Our SEETrials system demonstrated high precision, recall (sensitivity), and F1 scores, achieving overall metrics of 0.958, 0.944, and 0.951, respectively, across all trial phases. Supplement Table 2 displays the average performance scores for both the overall dataset and each phase of the MM studies.

We implemented our system on clinical trial reports from four other randomly chosen cancer types, including both solid cancers (lung and breast) and hematologic cancers (lymphoma and leukemia). The average precision, recall, and F1 scores were 0. 978, 0.969, 0.974 (breast cancer), 0.986, 0.966, 0.976 (lung cancer), and 0.981, 0.966, 0.974 (leukemia/lymphoma), respectively. Comprehensive details, including strict and relaxed evaluation scores, are provided in Supplement Table 3.

### Qualitative Error Analysis

We conducted a qualitative error analysis on individual documents. A prevalent inaccuracy in the extraction process pertained to cohort/arm identification and categorization. Abstracts frequently encompass diverse clinical information for multiple cohorts/arms, including dose-escalation, dose-expansion, recommended phase 2 dosage (RP2D) groups, entire groups, and subgroups. The lack of explicit descriptions in the text occasionally posed a challenge for the system in determining the appropriate allocation of values. Supplement Table 4 provides illustrative example findings and detailed descriptions. Additional errors included extracting the author’s affiliation as the study location in PubMed abstracts, cohort size discrepancies, and occasional extraction of only percentage values without other relevant information. Improvements are needed for consistent value extraction and allocation.

### Analyzing Efficacy and Safety Data from Abstracts

We extracted data from 130 abstracts related to CAR-T, 63 to BsAbs, 38 to ADC, 6 to CELMoD, 4 to HDACi, and 4 to ICI therapies. Among these, there were 75, 47, 46, and 16 abstracts corresponding to phases 1, 1/2, 2, and 3. 61 abstracts did not have phase information. The distribution of clinical trial phases in each therapy is detailed in Supplement Table 1. The complete list of extracted data elements from all studies is detailed in Supplement Figure 1.

Figure 2 visually summarizes frequently appearing efficacy and safety-related entity percentages across the three therapies, CAR-T, BsAbs, and ADC. We present a comprehensive breakdown of trial numbers and the percentages of each efficacy-related and safety-related entity out of all mentioned entities (Figures 3 and 4, respectively) categorized by clinical trial phases (phases 1, 1/2, 2, and 3) for both efficacy and safety-related entities.

**Figure 2.**
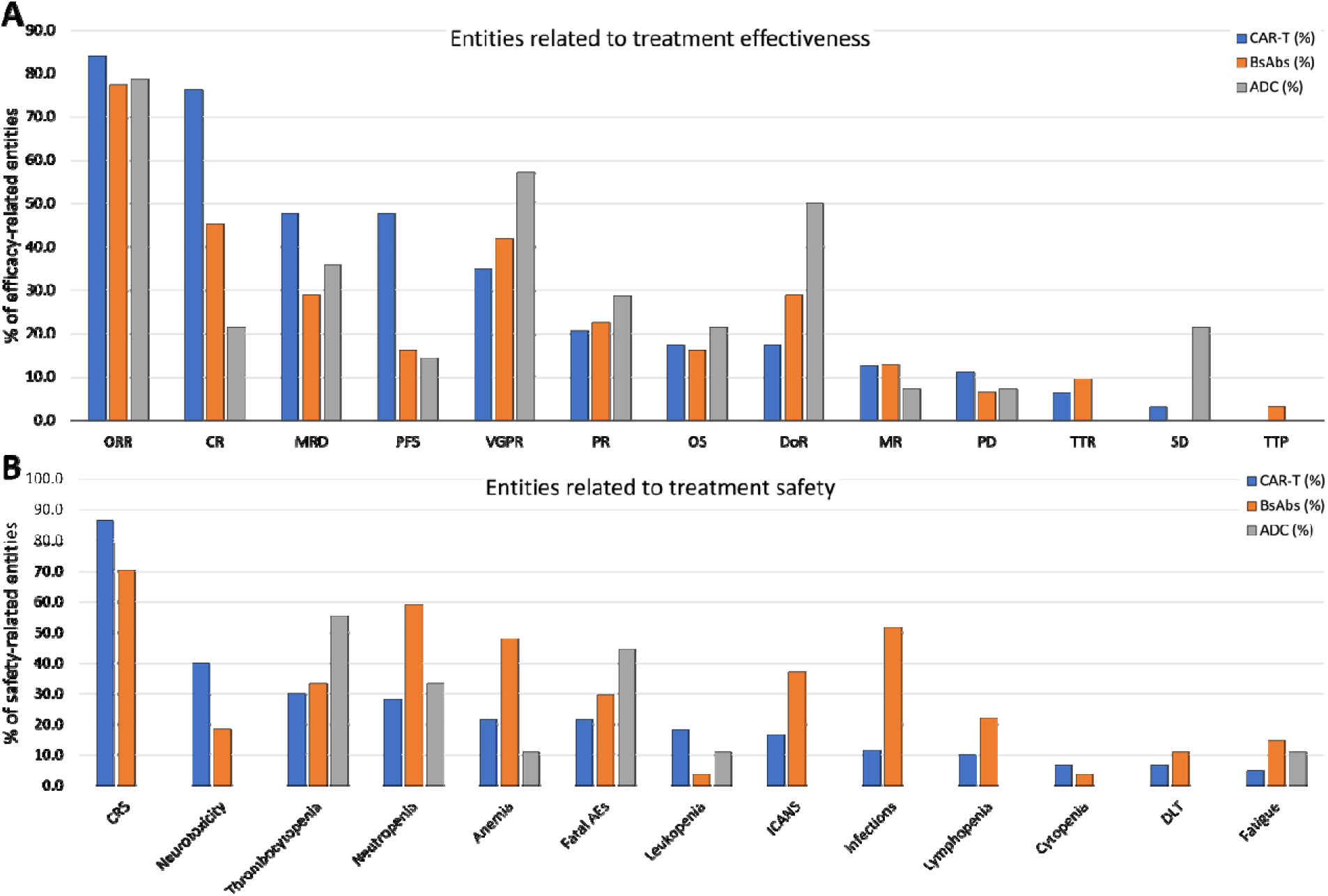
The comparative landscape of efficacy and safety entities across CAR-T, BsAbs, and ADC therapies. This visual summary illustrates the percentages of 11 efficacy and 13 safety-related entities across CAR-T cell therapy, BsAbs, and ADC therapies, providing a comprehensive overview of their comparative clinical profiles. A) Entities related to treatment effectiveness. B) Entities related to treatment safety. CAR-T, chimeric antigen receptor T cell; BsAbs, Bispecific antibody; ADC, antibody-drug conjugate; ORR, overall response rate; CR, complete response; VGPR, very good partial response; PR, partial response; PFS, progression-free survival; MRD, minimum residual disease; OS, overall survivor; DoR, duration of response; SD, stable disease; PD, progressive disease; TTR, time to response; MR, minimal response; TTP, time to progress; TTTD, time to treatment discontinuation; TTTF, time to treatment failure; TTNT, time to next treatment; DCR, disease control rate; CRS, cytokine release syndrome; Aes, adverse events; ICANS, immune effector cell associated neurotoxicity syndrome.

**Figure 3.**
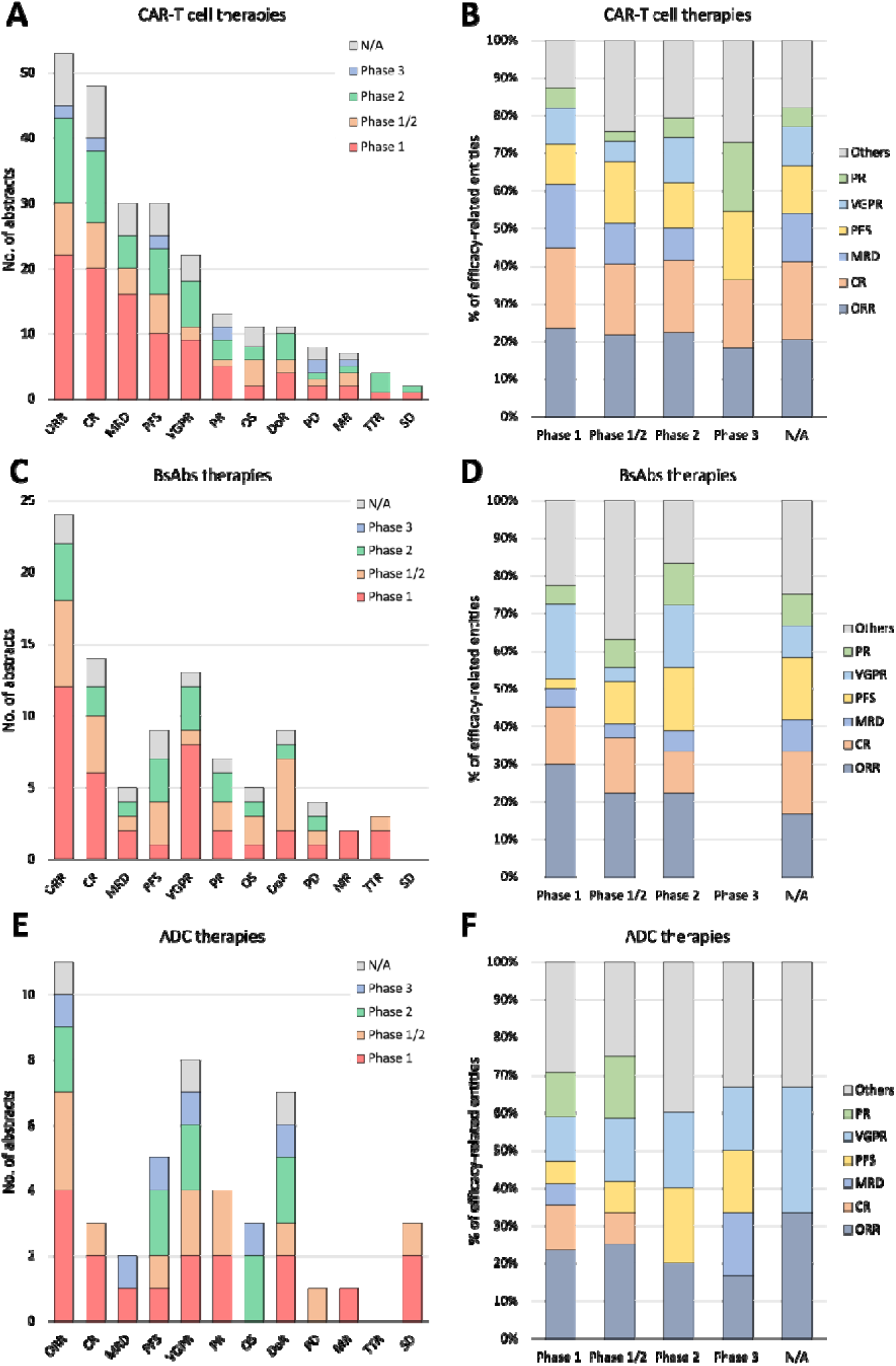
A detailed breakdown of abstract numbers with each efficacy-related entity (A, C, E) and percentages of each entity out of all mentioned entities (B, D, F) is presented, categorizing clinical trials into phases 1, 1/2, 2, and 3. A and B: CAR-T cell therapies. C and D: BsAbs therapies. E and F: ADC therapies. CAR-T, chimeric antigen receptor T cell; BsAbs, Bispecific antibody; ADC, antibody-drug conjugate; ORR, overall response rate; CR, complete response; (VG)PR, (very good) partial response; PFS, progression-free survival; MRD, minimum residual disease; OS, overall survivor; DoR, duration of response; SD, stable disease; PD, progressive disease; TTR, time to response; MR, minimal response.

**Figure 4.**
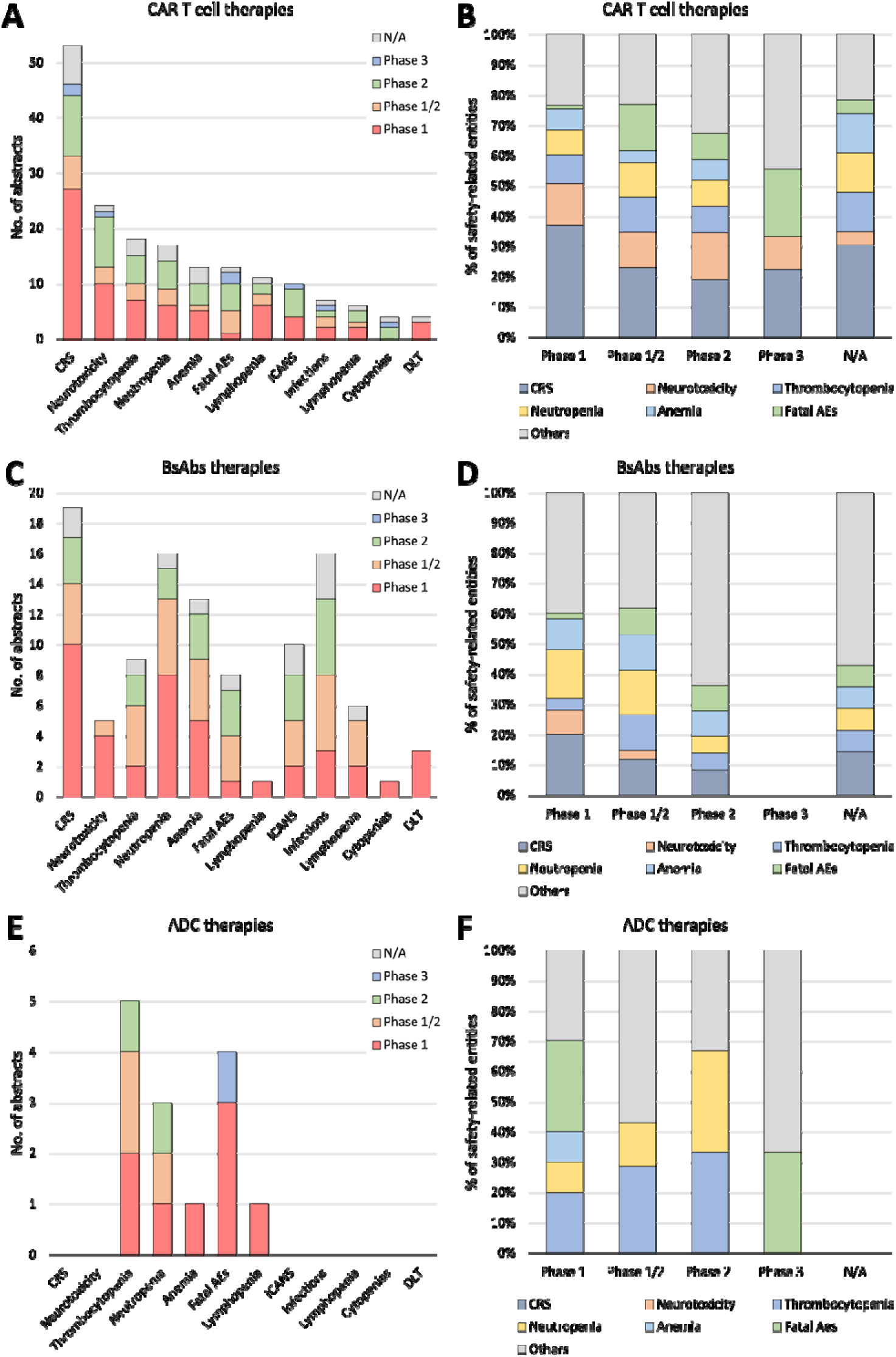
A detailed breakdown of abstract numbers with each safety-related entity (A, C, E) and percentages of each entity out of all mentioned entities (B, D, F) is presented, categorizing clinical trials into phases 1, 1/2, 2, and 3. A and B: CAR-T cell therapies. C and D: BsAbs therapies. E and F: ADC therapies. CAR-T, chimeric antigen receptor T cell; BsAbs, Bispecific antibody; ADC, antibody-drug conjugate; CRS, cytokine release syndrome; Fatal AEs, fatal adverse events; ICANS, immune effector cell associated neurotoxicity syndrome; DLT, Dose Limiting Toxicity.

The ORR emerged as the most frequently reported entity in clinical trial abstracts, constituting 84.1% in CAR-T, 77.4% in BsAbs, and 78.6% in ADC studies. CR was mentioned in 76.2% of CAR-T and 45.2% of BsAbs abstracts. Very good partial responses (VGPR) were mentioned in 57.1% of ADC and 41.9% of BsAbs abstracts (Supplement Table 5). Supplement Table 6 provides the percentage of abstracts mentioning each safety-related entity, categorized by phases and therapies. For ORR, 41.5%, 50.0%, and 36.4% were from phase 1 studies, and 15.1%, 25.0%, and 27.3% were from phase 1/2 studies for CAR-T, BsAbs, and ADC, respectively.

Similarly, for VGPR, 40.9%, 61.5%, and 25.0% were from phase 1 studies, and 31.8%, 23.1%, and 25% were from phase 2 studies for CAR-T, BsAbs, and ADC, respectively. In safety-related entity analysis, cytokine release syndrome (CRS) was prominently mentioned in CAR-T (86.7% of abstracts) and BsAbs (70.4% of abstracts) studies but not in ADC studies (0%). Infection was frequently cited in BsAbs (51.9%) compared to CAR-T (11.7%) and ADC (0%) abstracts. Conversely, thrombocytopenia was more commonly mentioned in ADC (55.6%), compared to CAR-T (30%) and BsAbs (33.3%). Notably, Fatal adverse events (AEs) or Severe AEs more prevalently appeared in ADC (44.4% and 44.4%, respectively) abstracts compared to CAR-T (21.7% and 3.3%) and BsAbs (29.6% and 7.4%) (Supplement Table 7). When assessing the percentage of abstracts mentioning each safety-related entity by phases and therapies (Supplement Table 8), our data revealed that most safety-related entities such as CRS, neurotoxicity, and thrombocytopenia were mentioned more frequently in phase 1 studies compared to phase 1/2 or phase 2 studies, except for fatal AEs in CAR-T (7.7%, 30.8%, and 38.5%, respectively) and BsAbs (12.5%, 37.5%, and 37.5%, respectively).

For the comparative analysis of treatment outcome values among CAR-T, BsAbs, and ADC therapies, we included a total of 93 distinct studies identified using NCT IDs. For studies lacking NCT IDs, we manually examined the authors, their affiliations, and study titles. Of these, 87 were included in the efficacy comparison and 74 were included in the safety comparison. Supplement Table 9 provides a detailed breakdown of the trial numbers by therapies and phases. The degrees of study heterogeneity for ORR (*I^2^*=87%, 97%, and 96%, respectively) (Supplement Figure 2A) and CR (*I^2^*=90%, 91%, and 94%, respectively) (Figure 4B) were high (over 75%) in all three therapies even after categorized by phases for ORR (Phase 1: *I^2^*=81%, 76%, and 85%; Phase 2: *I^2^*=88%, 97%, and 95%, respectively) (Figure 5 A and B) and for CR (Phase 1: *I^2^*=87%, 39%, and 96%; Phase 2: *I^2^*=91%, 96%, and NA, respectively) (Figure 5 C and D) except phase 1 BsAbs studies (*I^2^*=39%). The 95% CIs of estimated ORR and CR were 84-92% and 53-66% (CAR-T), 55-73% and 16-37% (BsAbs), and 37-65% and 1-51% (ADC). We conducted a similar analysis on CRS (including studies mentioning all grades but excluding studies only mentioning grade >=3) and Neutropenia grade >=3 for each therapy. The 95% CIs of estimated CRS and grade >=3 neutropenia were 64-86% and 65-93% (CAR-T), 51-74% and 38-55% (BsAbs), and N/A and 2-29% (ADC). The results were visualized in Supplement Figure 3 (overall) and Figure 6 (for phases 1 and 2).

**Figure 5.**
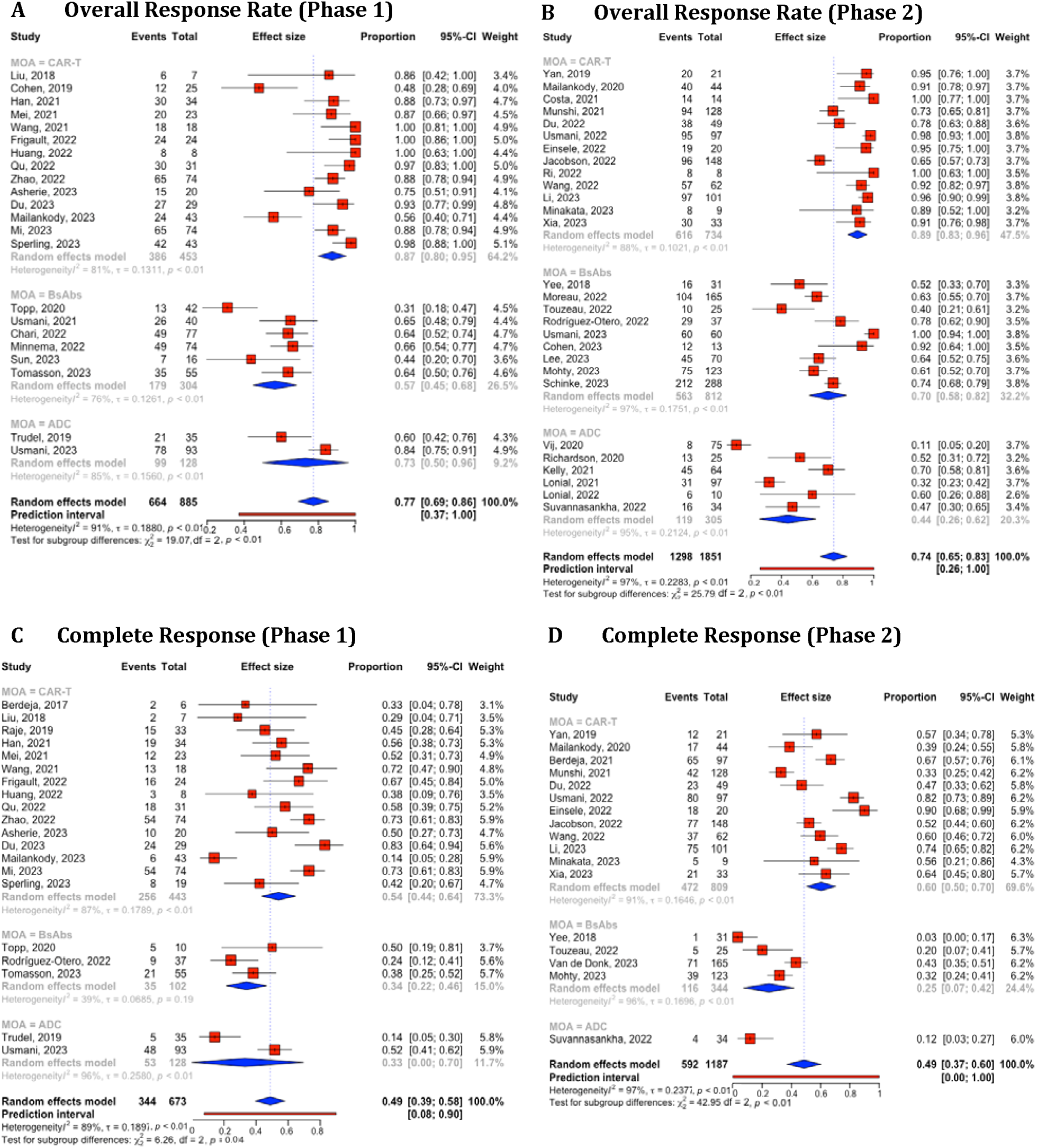
Combined and subgroup analysis of overall response rate and complete response based on therapies categorized in phase 1 and 2 trials. A. Overall response rates in phase 1 trial. B. Overall response rates in phase 2 trial. C. Complete response in phase 1 trial. D. Complete response in phase 2 trial. Horizontal lines through the squares indicate 95% Confidence Intervals (CIs). The diamond symbol aggregates these estimates, presenting the pooled mean effect size and its 95% CI. CAR-T, chimeric antigen receptor T cell; BsAbs, Bispecific antibody; ADC, antibody-drug conjugate.

**Figure 6.**
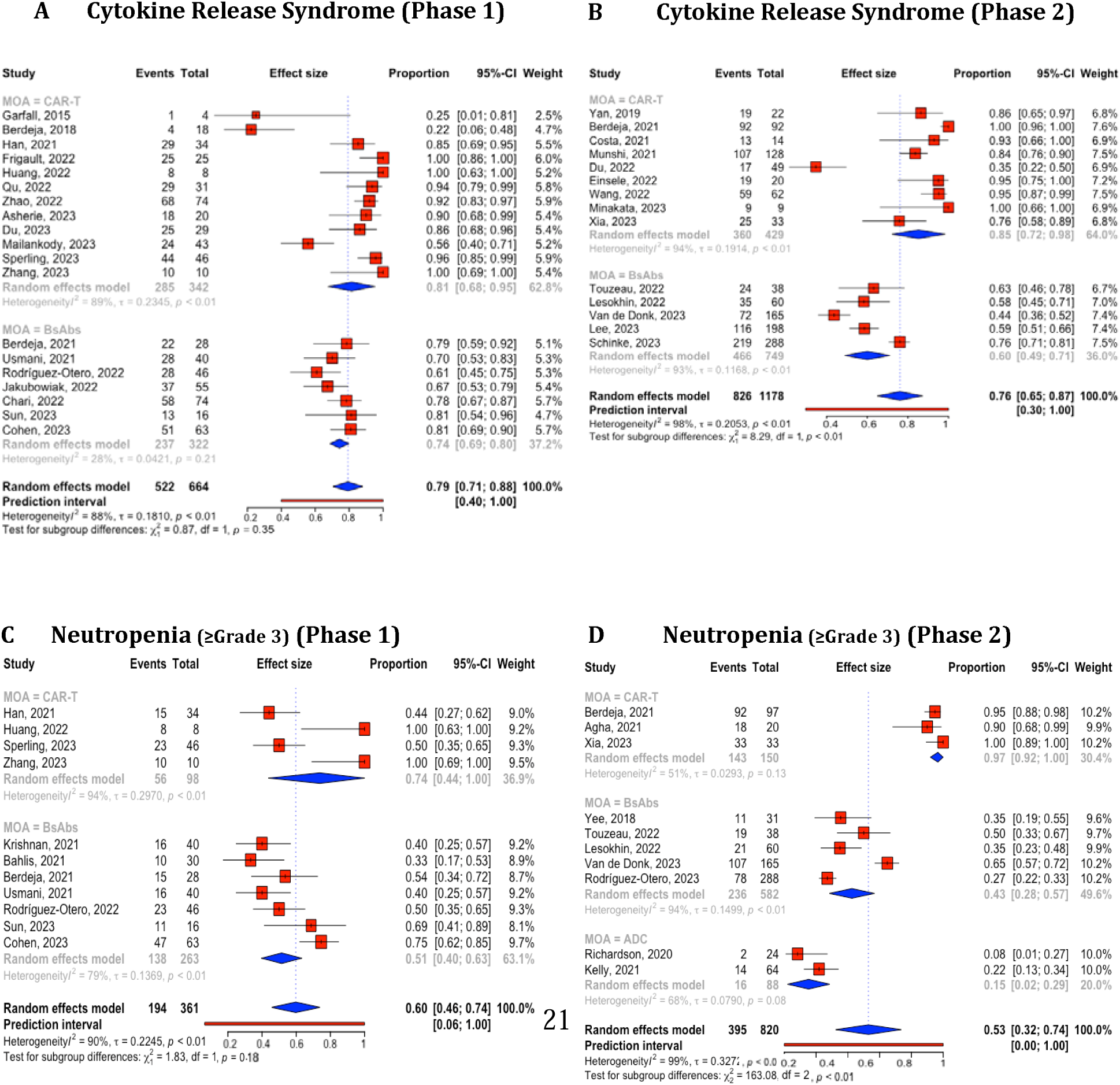
Combined and subgroup analysis of cytokine release syndrome and neutropenia (≥Gr3) based on therapies categorized in phase 1 and 2 trials. A. Cytokine release syndrome in phase 1 trial. B. Cytokine release syndrome in phase 2 trial. C. Neutropenia (≥Gr3) in phase 1 trial. D. Neutropenia (≥Gr3) in phase 2 trial. Horizontal lines through the squares indicate 95% Confidence Intervals (CIs). The diamond symbol aggregates these estimates, presenting the pooled mean effect size and its 95% CI.CAR-T, chimeric antigen receptor T cell; BsAbs, Bispecific antibody; ADC, antibody-drug conjugate.

## DISCUSSION

Our knowledge-driven LLM system, SEETrials, demonstrated the capabilities of GPT-4 in extracting and processing clinical trial data. The success of SEETrials in automatically identifying intervention outcomes and their associations with specific cohorts is underscored by robust performance metrics and speaks to the potential of LLMs to revolutionize data extraction from condensed and complex medical texts.

The system’s proficiency in handling data from annual conference abstracts is particularly noteworthy. These abstracts are often the first public report of a clinical trial’s findings and can influence clinical practice ahead of peer-reviewed publications. The ability of SEETrials to process this information rapidly and reliably is invaluable, ensuring that the latest clinical insights are accessible to healthcare professionals and researchers without delay. It also showcased the generalizability of a prompt initially tailored for specific cancer (i.e., multiple myeloma) to effectively extract trial information from diverse cancer types (breast cancer, lung cancer, and leukemia/lymphoma) with consistently high-performance scores across various disease clinical trial studies. This enables uniform analysis of clinical trial outcomes across diseases. To our knowledge, this study pioneers exploring GPT-powered models for comprehensive clinical trial outcomes extraction from diverse abstract types, in addition to our previously established system “AutoCriteria”—a generalizable clinical trial eligibility criteria extraction system powered by a large language model ^24^.

Importantly, our study extends beyond mere data extraction. By conducting a comparative analysis of CAR-T, BsAbs, and ADC therapies specifically within MM clinical trials, we have gleaned insights into the varying safety and efficacy profiles across different treatment modalities and trial phases. The predominance of certain efficacy markers, such as ORR, in phase 1 trials underscores the importance of these early-phase trials in gauging initial treatment impact. Conversely, the emphasis on survival and treatment duration in phase 2 trials highlights the shift towards understanding the long-term benefits and potential risks of therapies as they progress through the clinical trial pipeline. Our findings also align with existing literature, revealing consistent safety profiles in CAR-T, BsAbs, and ADC studies such as higher CRS rates in CAR-T therapy and infection rates in ADCs ^26, 27^. Understanding these outcomes’ variations across therapies and phases is crucial for contextualizing clinical trial results.

Moreover, our findings highlight a concerning trend: the increase in reports of fatal adverse events in later trial phases, particularly for CAR-T and ADC therapies. It suggests that early-phase studies may not fully capture the adverse event profile, especially for treatments with complex mechanisms of action or those applied to severely ill patient populations. This observation calls for a re-evaluation of safety monitoring protocols, especially as therapies advance beyond the dose-escalation stages. We also observed a higher number of abstracts mentioning fatal or severe AEs entities in ADC studies compared to CAR-T and BsAbs. We noted significant variability in the effectiveness of the same treatments, even when categorized by phases, for outcomes such as Objective Response Rate (ORR), Complete Response (CR), Cytokine Release Syndrome (CRS), and neutropenia. A possible reason for the exceptionally low ORR observed in some Phase 1 studies may be attributed to combining data from different dosage groups, as indicated in the abstracts.

We recognize several limitations in our study. The exclusive focus on abstracts, while practical, limits the depth of our analysis. Critical nuances contained within full-text articles are often lost in abstracts, which could lead to incomplete or skewed understandings of the safety and efficacy profiles of therapies. Additionally, the omission of details such as prior treatment histories, specific high-risk patient groups, and biomarker statuses from our analysis may have prevented a fully informed assessment of the treatments’ impacts.

The future potential of SEETrials is promising. Extending the system’s capabilities to incorporate full-text articles, detailed patient histories, and biomarker data could enrich the dataset, leading to even more comprehensive insights. A comparison of outcomes between preliminary conference abstracts and subsequent full publications could also be instrumental in evaluating the consistency and progression of reported data ^28^. Such an expansion could serve as a valuable tool for the medical community, addressing key inquiries and shaping clinical decision-making processes. Furthermore, the integration of SEETrials’ extracted results with expert-curated databases like HemOnc could pave the way for an augmented, collaborative approach to data analysis in oncology, ensuring that the most relevant and accurate information is available to inform patient care and research endeavors ^29^.

## CONCLUSION

We developed SEETrials, an LLM-based system with broad applicability, capable of identifying granular safety and efficacy entities across various clinical trial studies. SEETrials shows significant potential in accurate data extraction and comparison, facilitating efficient and timely generation and dissemination of clinical evidence.

## AUTHOR CONTRIBUTIONS

KL, HP, and XW conceptualized the study design and spearheaded the development and evaluation of the system. HP, KL, LH, and SD led the data extraction, prompt engineering, and computational analysis, and drafted the initial manuscript. KL, HP, and NO oversaw the knowledge ingestion, gold standard and coordinated the evaluation of the system. KL, HP, SD, LH, FM, BH, SMR, AJC, MK, JLW, HX, and XW edited and revised the manuscript. LH, JH, and JW provided the infrastructure support for computation. All authors read and approved the final manuscript.

## DATA AVAILABILITY

The SEETrials source code, prompts, and output are available at https://github.com/applebyboy/SEEtrials.git

---

**Supplement Table 1.**
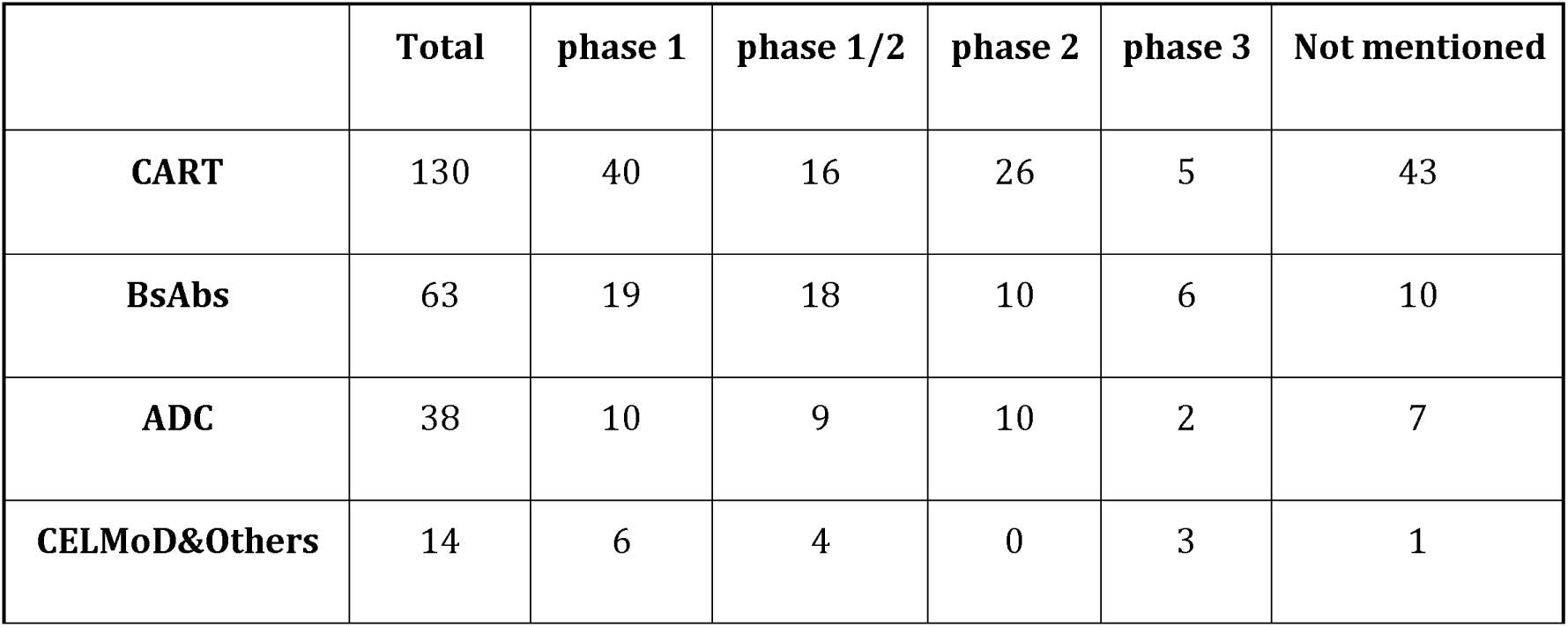
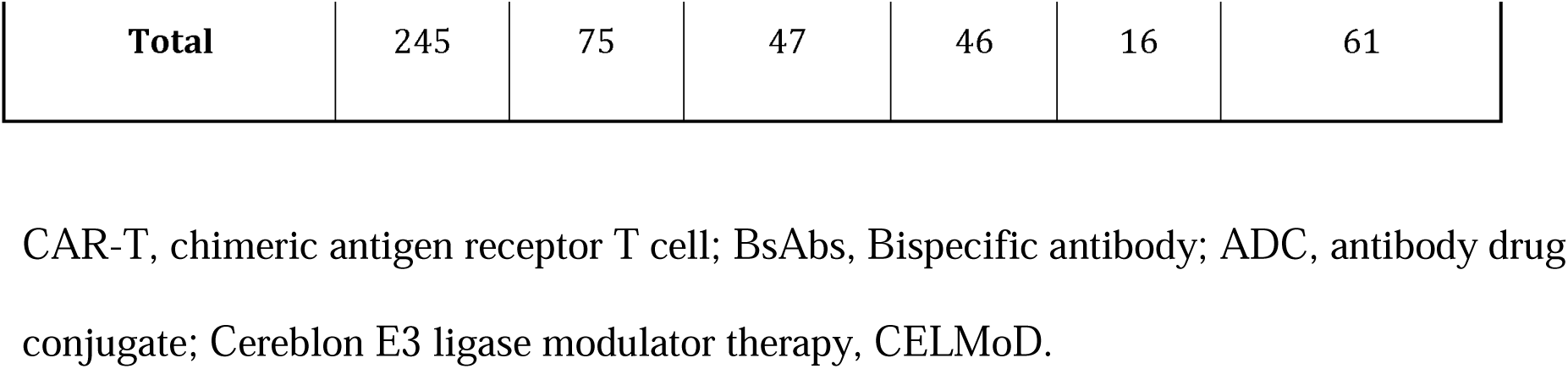
Characteristics overview of abstracts included.

|  | Total | phase 1 | phase 1/2 | phase 2 | phase 3 | Not mentioned |
| --- | --- | --- | --- | --- | --- | --- |
| **CART** | 130 | 40 | 16 | 26 | 5 | 43 |
| **BsAbs** | 63 | 19 | 18 | 10 | 6 | 10 |
| **ADC** | 38 | 10 | 9 | 10 | 2 | 7 |
| **CELMoD&Others** | 14 | 6 | 4 | 0 | 3 | 1 |
| **Total** | 245 | 75 | 47 | 46 | 16 | 61 |

CAR-T, chimeric antigen receptor T cell; BsAbs, Bispecific antibody; ADC, antibody drug conjugate; Cereblon E3 ligase modulator therapy, CELMoD.

**Supplement Table 2.** Performance metrics of the SEETrials system in multiple myeloma clinical trial study abstracts.

| Phase | No. of Abstracts | Precision | Recall | F1-score |
| --- | --- | --- | --- | --- |
| 1 | 36 | 0.939 | 0.929 | 0.934 |
| 1/2 | 23 | 0.982 | 0.970 | 0.976 |
| 2 | 17 | 0.976 | 0.963 | 0.969 |
| 3 | 7 | 0.948 | 0.936 | 0.942 |
| N/A | 17 | 0.947 | 0.923 | 0.935 |
| **Total** | **100** | **0.958** | **0.944** | **0.951** |

**Supplement Table 3.** Performance metrics of the SEETrials system in various cancer clinical trial study abstracts.

| Disease type | Abstract No. | Precision | Recall | F1-score |
| --- | --- | --- | --- | --- |
|  | 115 | 0.982 | 0.967 | 0.975 |
| Breast cancer | 30 | 0.978 | 0.969 | 0.974 |
| Leukemia/<br>Lymphoma | 50 | 0.981 | 0.966 | 0.974 |
| Lung cancer | 35 | 0.986 | 0.966 | 0.976 |

**Supplement Table 4.**
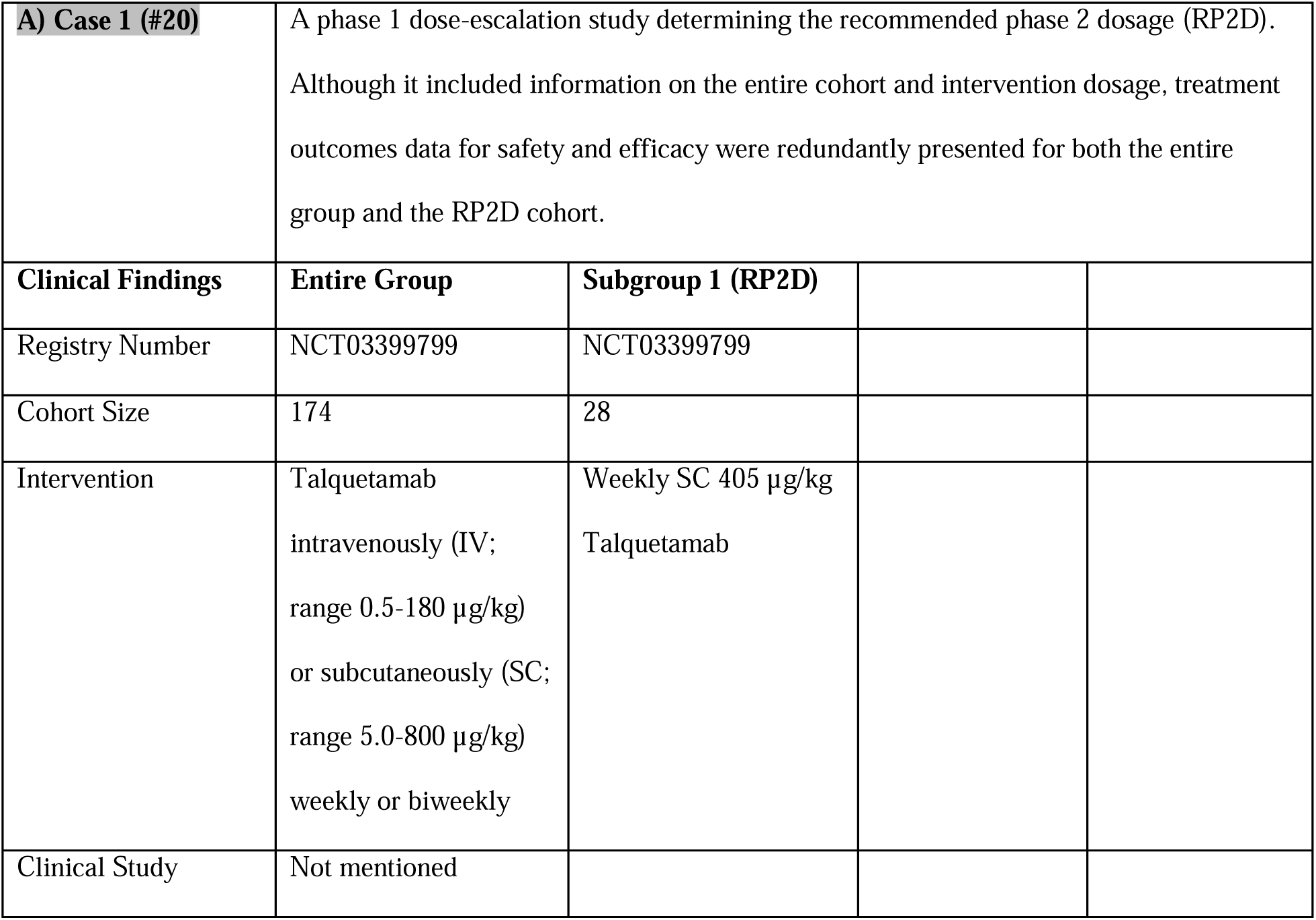
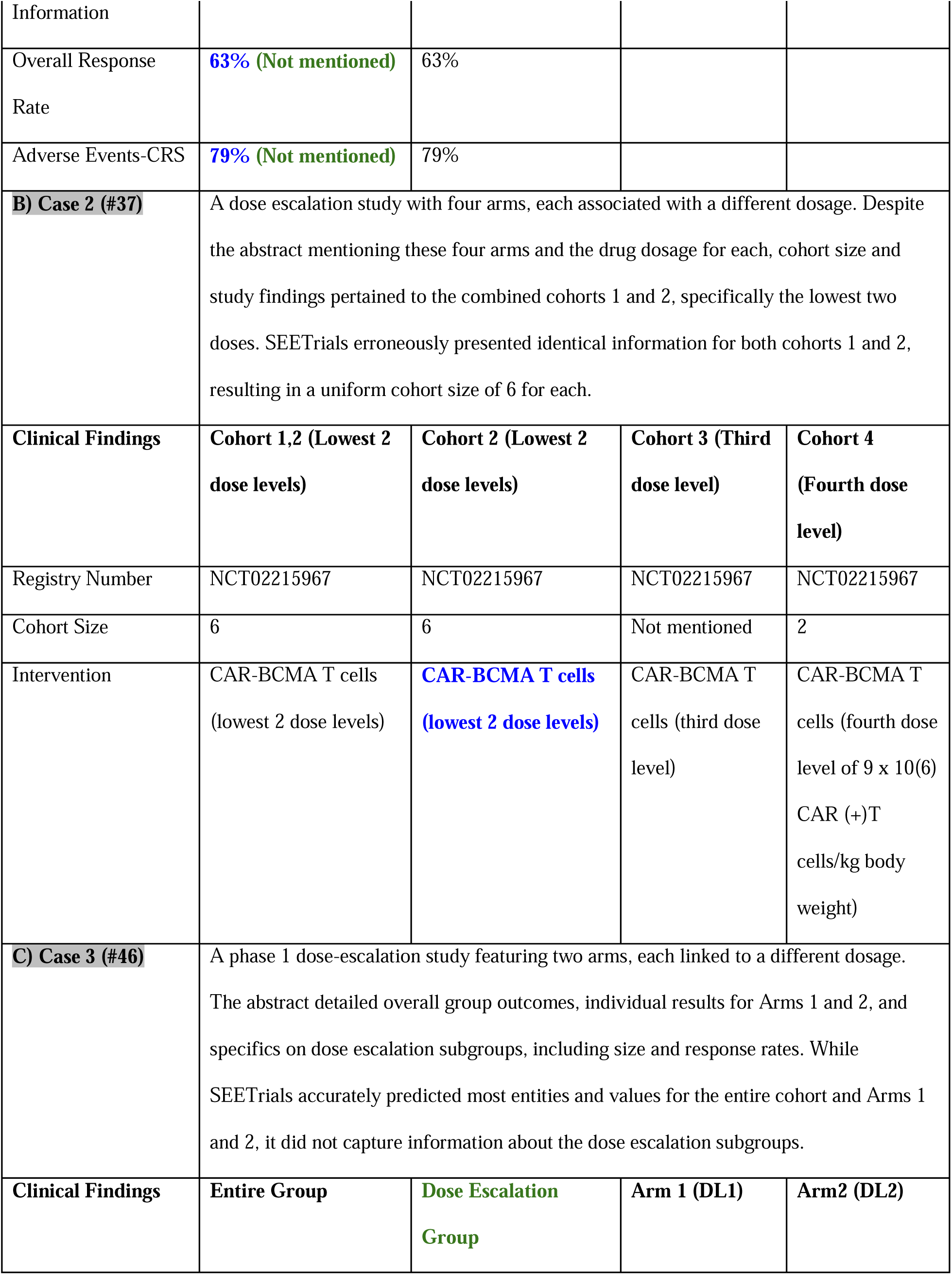
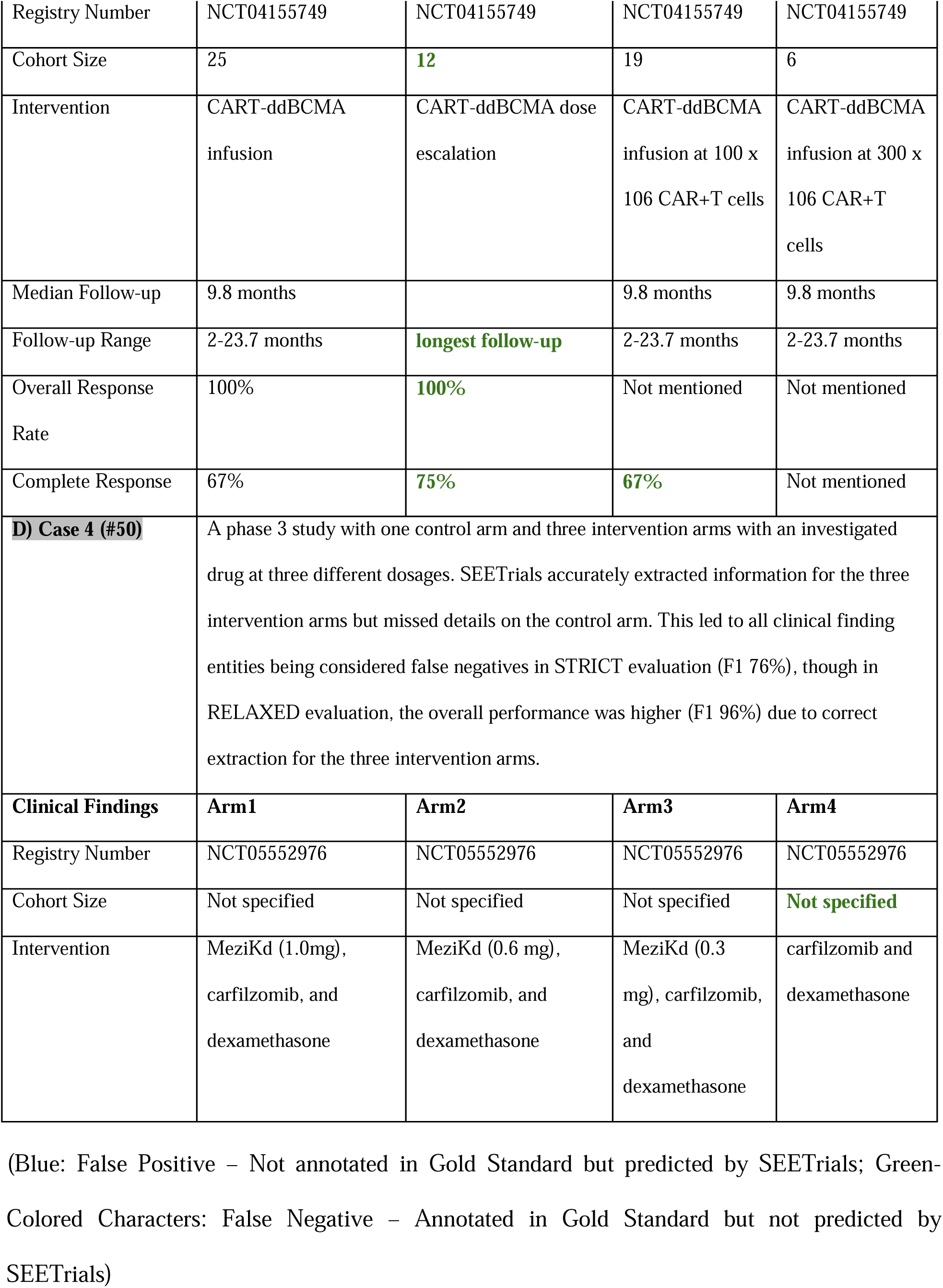
Examples of qualitative error analysis. A) Partial output from the abstract #20 ASCO_ADC. B) Partial output from abstract #37 PubMed_CAR-T. C) Partial output from abstract #46 ASCO_CAR-T. D) Partial output from abstract #50_ASCO_CELDoM.

**Supplement Table 5.**
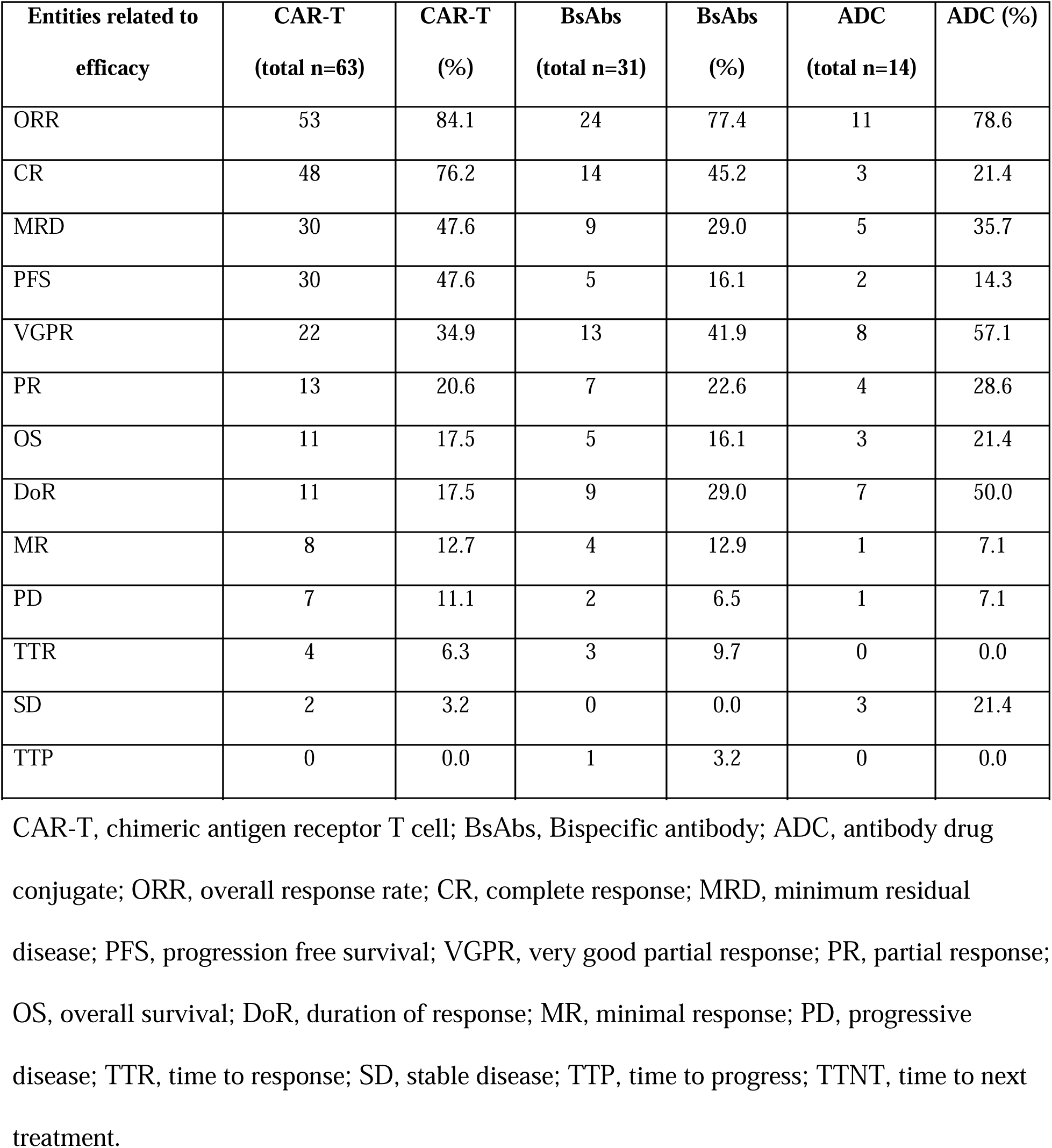
Descriptive statistics of extracted clinical outcomes on treatment efficacy. (N represents the absolute number of abstracts where each entity is mentioned, and the percentage (%) was calculated by dividing it by the total number of abstracts for each therapy)

| **Entities related to efficacy** | **CAR-T**<br>**(total n=63)** | **CAR-T**<br>**(%)** | **BsAbs**<br>**(total n=31)** | **BsAbs**<br>**(%)** | **ADC**<br>**(total n=14)** | **ADC (%)** |
| --- | --- | --- | --- | --- | --- | --- |
| ORR | 53 | 84.1 | 24 | 77.4 | 11 | 78.6 |
| CR | 48 | 76.2 | 14 | 45.2 | 3 | 21.4 |
| MRD | 30 | 47.6 | 9 | 29.0 | 5 | 35.7 |
| PFS | 30 | 47.6 | 5 | 16.1 | 2 | 14.3 |
| VGPR | 22 | 34.9 | 13 | 41.9 | 8 | 57.1 |
| PR | 13 | 20.6 | 7 | 22.6 | 4 | 28.6 |
| OS | 11 | 17.5 | 5 | 16.1 | 3 | 21.4 |
| DoR | 11 | 17.5 | 9 | 29.0 | 7 | 50.0 |
| MR | 8 | 12.7 | 4 | 12.9 | 1 | 7.1 |
| PD | 7 | 11.1 | 2 | 6.5 | 1 | 7.1 |
| TTR | 4 | 6.3 | 3 | 9.7 | 0 | 0.0 |
| SD | 2 | 3.2 | 0 | 0.0 | 3 | 21.4 |
| TTP | 0 | 0.0 | 1 | 3.2 | 0 | 0.0 |
| TTNT | 0 | 0.0 | 1 | 3.2 | 0 | 0.0 |

CAR-T, chimeric antigen receptor T cell; BsAbs, Bispecific antibody; ADC, antibody drug conjugate; ORR, overall response rate; CR, complete response; MRD, minimum residual disease; PFS, progression free survival; VGPR, very good partial response; PR, partial response; OS, overall survival; DoR, duration of response; MR, minimal response; PD, progressive disease; TTR, time to response; SD, stable disease; TTP, time to progress; TTNT, time to next treatment.

**Supplement Table 6.** The distribution of abstracts mentioned each efficacy-related entity stratified by phases. (The percentage was calculated by dividing the number of abstracts where each entity is mentioned in each phase by the total number of abstracts with each entity mentioned)

| **Clinical Outcome** | **ORR** | **CR** | **PFS** | **VGPR** | **MRD** | **PR** |
| --- | --- | --- | --- | --- | --- | --- |
| **CAR-T** | **100%** | **100%** | **100%** | **100%** | **100%** | **100%** |
| Phase 1 | 41.5% | 41.7% | 33.3% | 53.3% | 40.9% | 38.5% |
| Phase 1/2 | 15.1% | 14.6% | 20.0% | 13.3% | 9.1% | 7.7% |
| Phase 2 | 24.5% | 22.9% | 23.3% | 16.7% | 31.8% | 23.1% |
| Phase 3 | 3.8% | 4.2% | 6.7% | 0.0% | 0.0% | 15.4% |
| N/A | 15.1% | 16.7% | 16.7% | 16.7% | 18.2% | 15.4% |
| **BsAbs** | **100%** | **100%** | **100%** | **100%** | **100%** | **100%** |
| Phase 1 | 50.0% | 42.9% | 40.0% | 11.1% | 61.5% | 28.6% |
| Phase 1/2 | 25.0% | 28.6% | 20.0% | 33.3% | 7.7% | 28.6% |
| Phase 2 | 16.7% | 14.3% | 20.0% | 33.3% | 23.1% | 28.6% |
| Phase 3 | 0.0% | 0.0% | 0.0% | 0.0% | 0.0% | 0.0% |
| N/A | 8.3% | 14.3% | 20.0% | 22.2% | 7.7% | 14.3% |
| **ADC** | **100%** | **100%** | **100%** | **100%** | **100%** | **100%** |
| Phase 1 | 36.4% | 66.7% | 50.0% | 20.0% | 25.0% | 50.0% |
| Phase 1/2 | 27.3% | 33.3% | 0.0% | 20.0% | 25.0% | 50.0% |
| Phase 2 | 18.2% | 0.0% | 0.0% | 40.0% | 25.0% | 0.0% |
| Phase 3 | 9.1% | 0.0% | 50.0% | 20.0% | 12.5% | 0.0% |
| N/A | 9.1% | 0.0% | 0.0% | 0.0% | 12.5% | 0.0% |

CAR-T; chimeric antigen receptor T cell; BsAbs, Bispecific antibody; ADC, antibody drug conjugate; ORR, overall response rate; CR, complete response; PFS, progression free survival; VGPR, very good partial response; MRD, minimum residual disease; PR, partial response

**Supplement Table 7.**
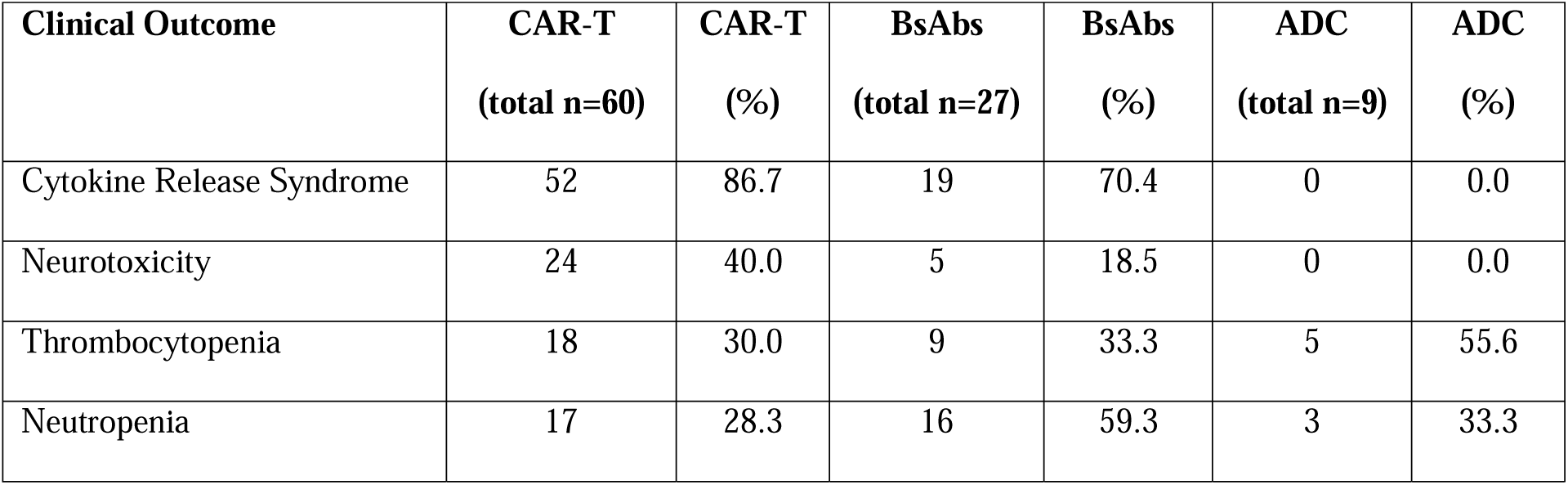
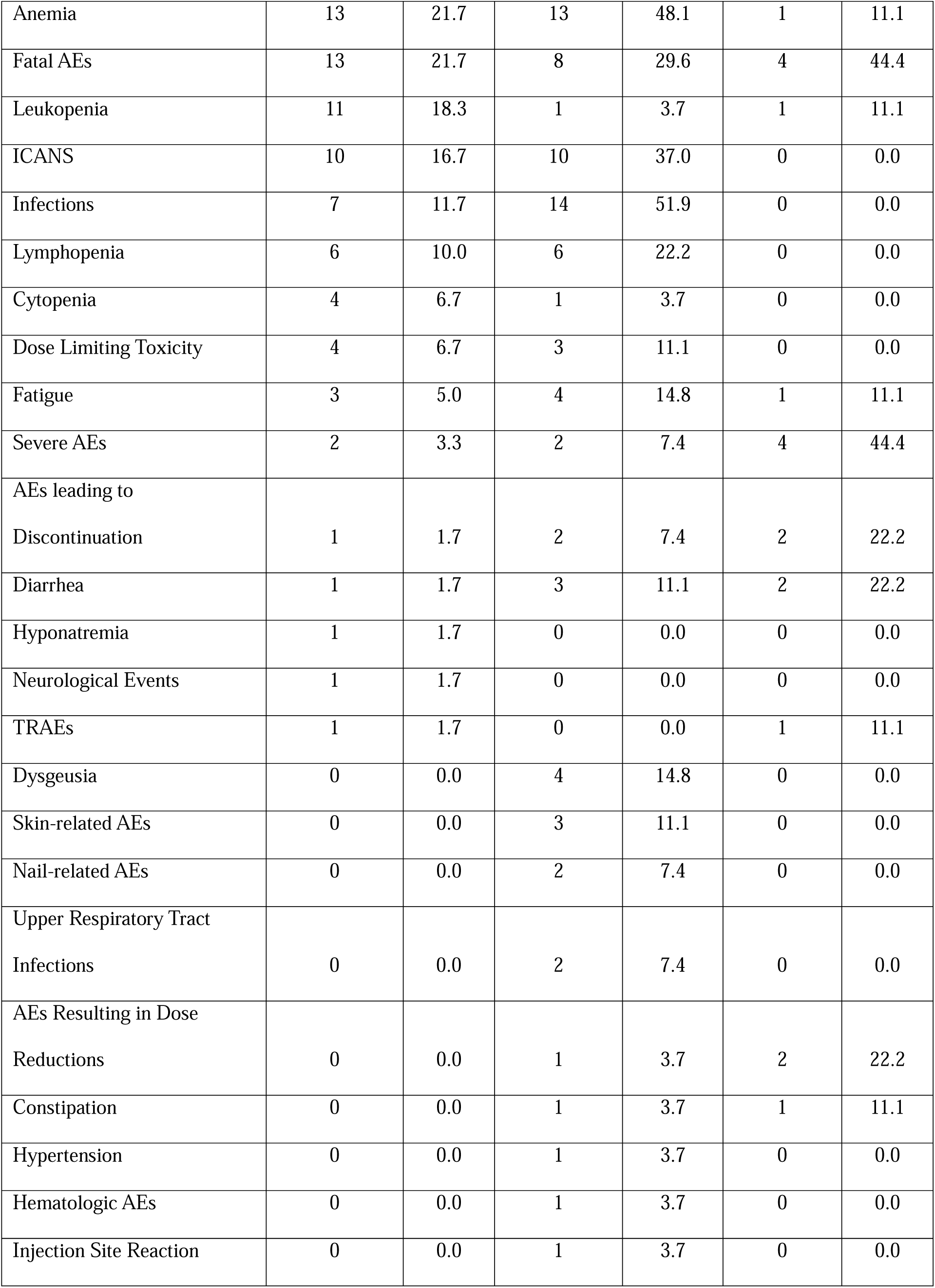
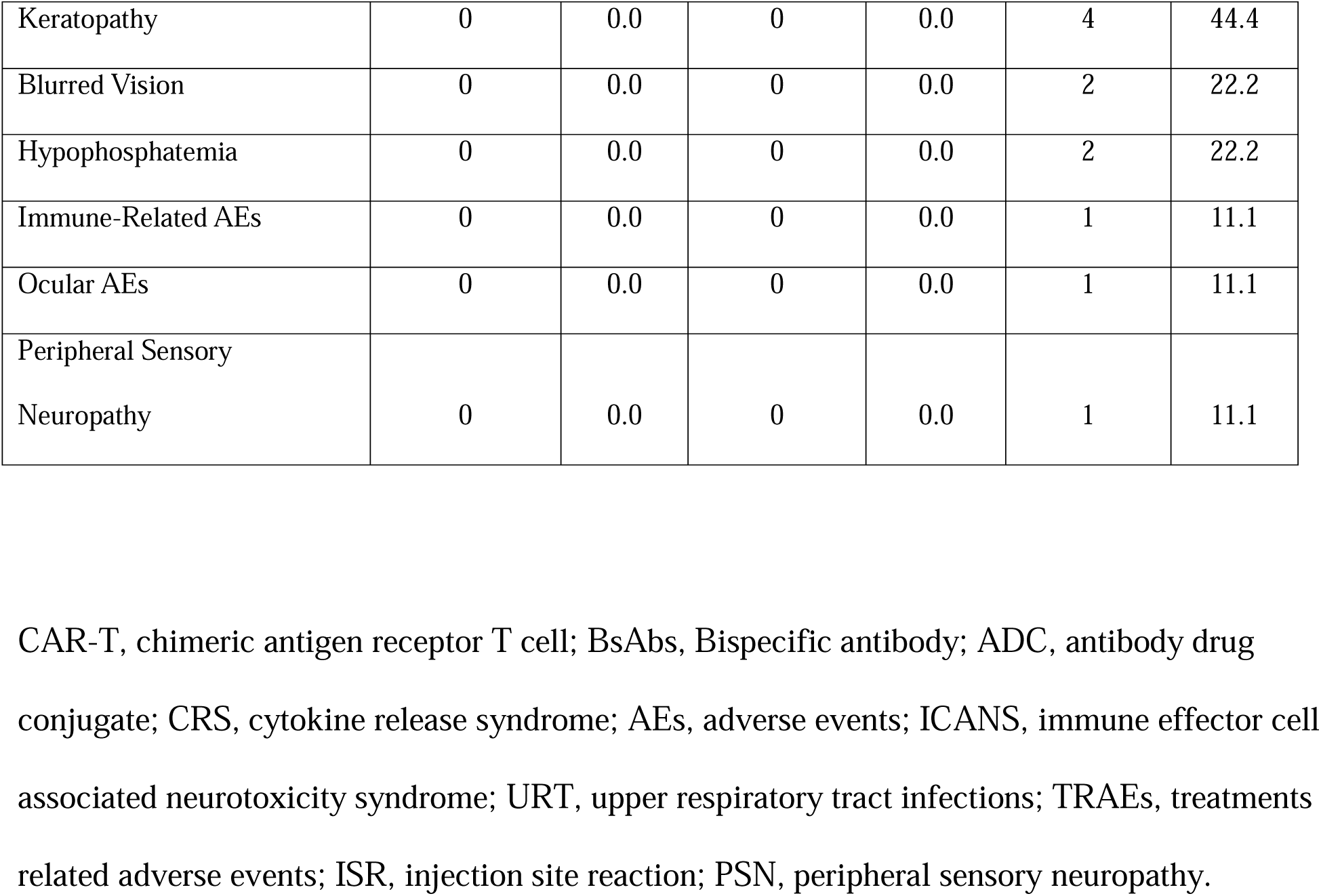
Descriptive statistics of extracted clinical outcomes on treatment safety. (N represents the absolute number of abstracts where each entity is mentioned, and the percentage (%) was calculated by dividing it by the total number of abstracts with safety outcome mentioned for each therapy)

**Supplement Table 8.**
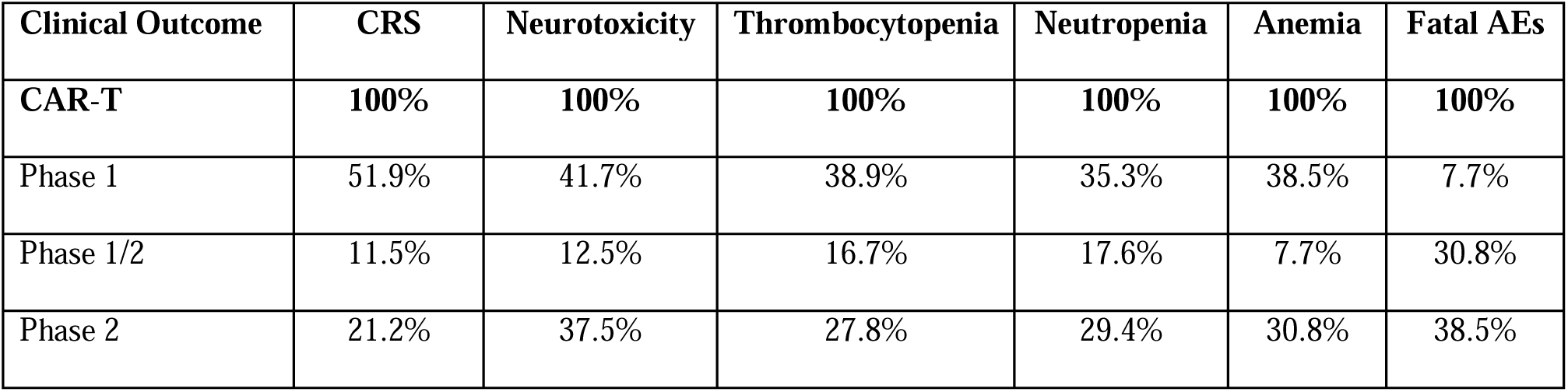
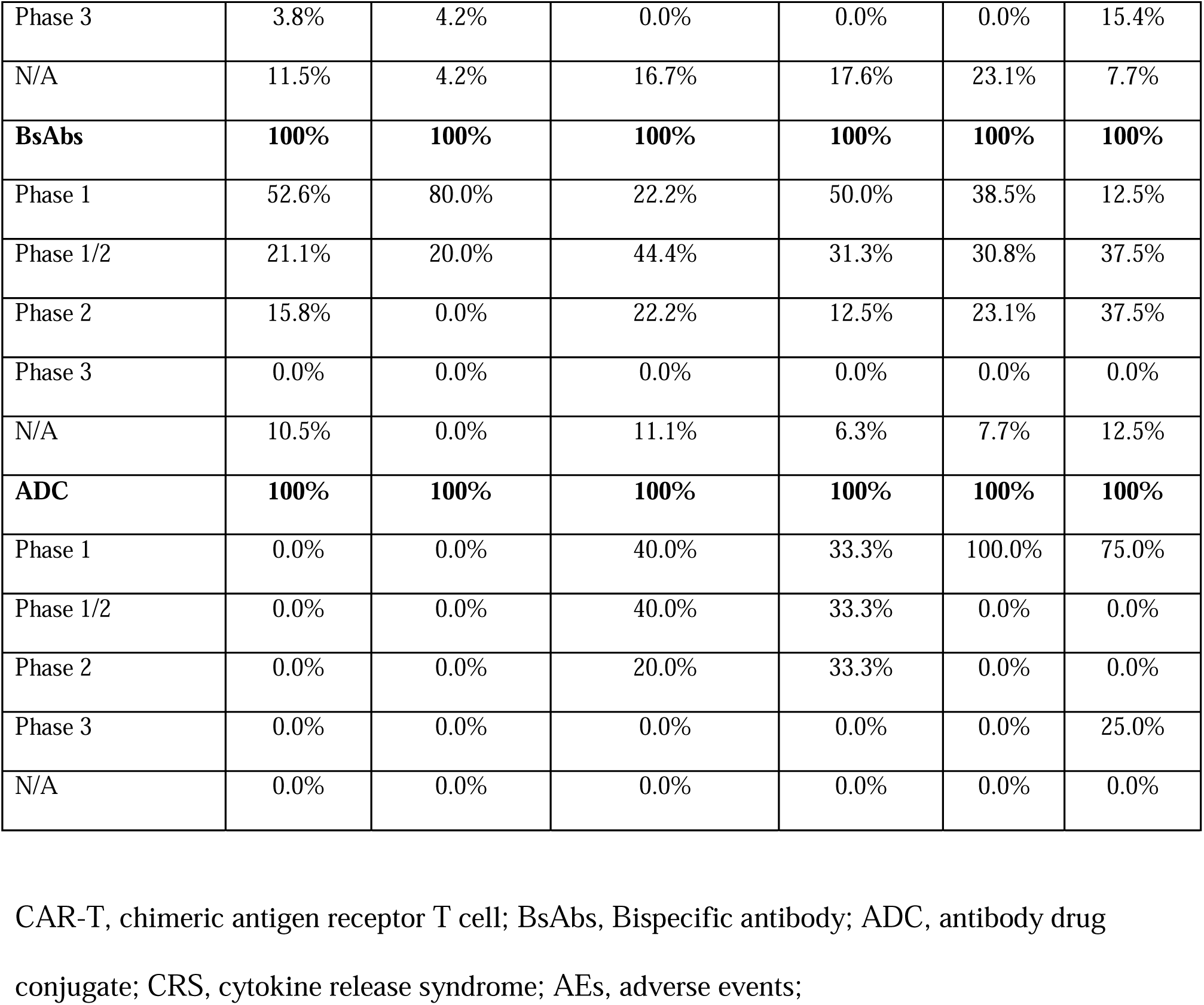
The distribution of abstracts mentioned each safety-related entity stratified by phases. (The percentage was calculated by dividing the number of abstracts where each entity is mentioned in each phase by the total number of abstracts with each entity mentioned)

**Supplement Table 9.** Detailed breakdown of the trial numbers utilized in comparative analysis, categorized by stages.

| **Efficacy** | **Total** | **phase 1** | **phase 1/2** | **phase 2** | **phase 3** | **Not mentioned** |
| --- | --- | --- | --- | --- | --- | --- |
| **CART** | 52 | 24 | 8 | 12 | 2 | 6 |
| **BsAbs** | 24 | 13 | 6 | 4 | 0 | 1 |
| **ADC** | 11 | 3 | 4 | 2 | 1 | 1 |
| **Total** | 87 | 40 | 18 | 18 | 3 | 8 |
| **Safety** | **Total** | **phase 1** | **phase 1/2** | **phase 2** | **phase 3** | **Not mentioned** |
| **CART** | 51 | 22 | 6 | 12 | 2 | 9 |
| **BsAbs** | 21 | 10 | 5 | 4 | 0 | 2 |
| **ADC** | 2 | 0 | 1 | 1 | 0 | 0 |
| **Total** | 74 | 32 | 12 | 17 | 2 | 11 |

CAR-T, chimeric antigen receptor T cell; BsAbs, Bispecific antibody; ADC, antibody drug conjugate; CRS, cytokine release syndrome; AEs, adverse events;

**Supplement Figure 1.**
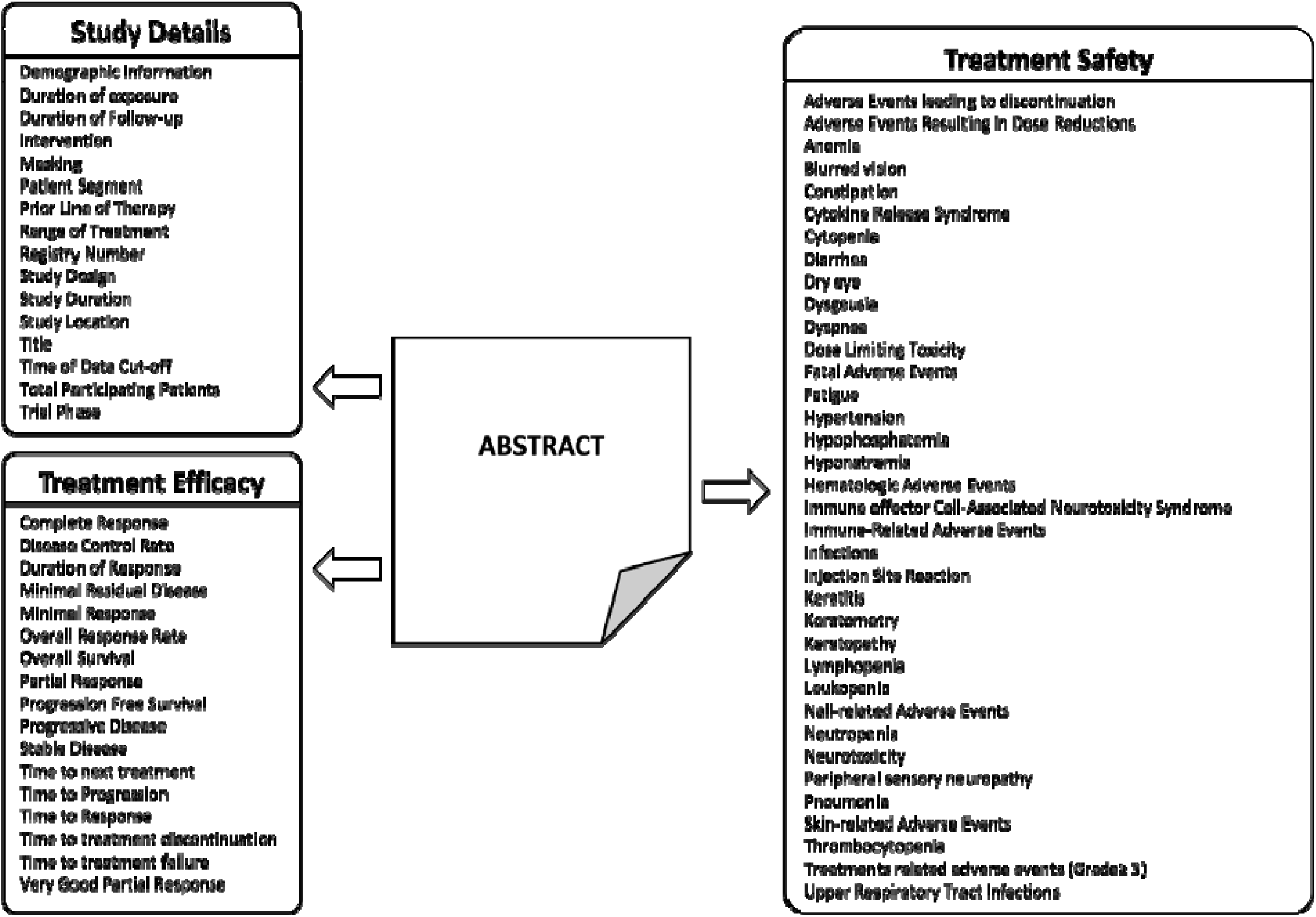
Comprehensive overview of extracted granular entities for study details, treatment efficacy, and safety. This illustrates a comprehensive breakdown of extracted granular entities from all multiple myeloma abstracts, encompassing Study Details, Treatment Efficacy, and Treatment Safety parameters, with a total of 16, 16, and 36 entities respectively, offering a detailed overview of the clinical trial landscape.

**Supplement Figure 2.**
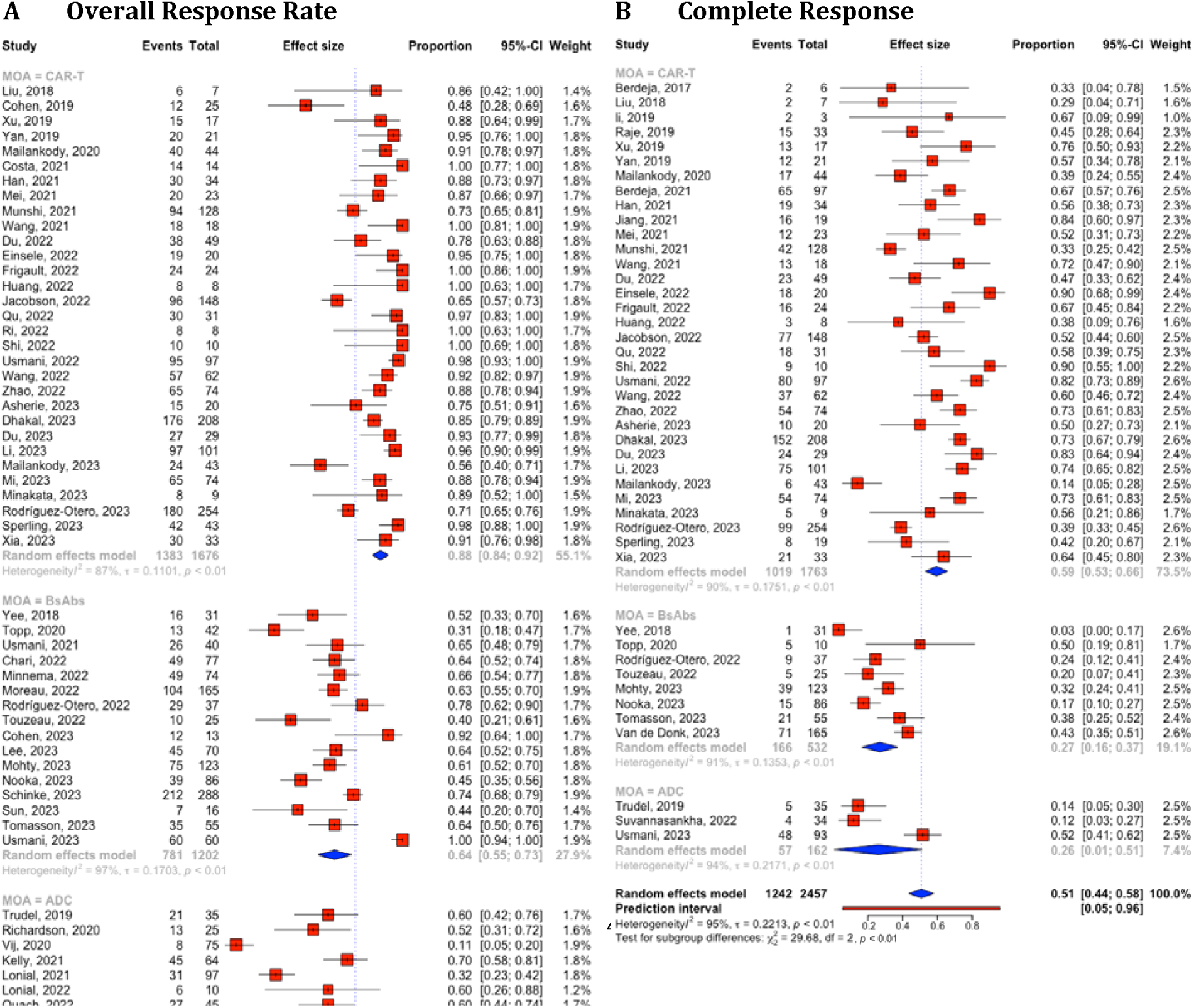
Combined quantitative analysis and subgroup analysis of overall response rate and complete response based on the therapies. A. Overall response rates. B. Complete response. The plot visualizes the estimated effect sizes from individual studies and their subgroups. Each square represents an estimated effect size, with the size proportional to the study’s weight in the analysis. Horizontal lines through the squares indicate 95% Confidence Intervals (CIs), offering insight into the precision of each study’s estimate. The diamond symbol aggregates these estimates, presenting the pooled mean effect size and its 95% CI, illustrating the overall effect across studies. The presence of a horizontal prediction line at the bottom of the plot provides a prediction interval, forecasting the range of effect sizes expected in similar future studies. CAR-T, chimeric antigen receptor T cell; BsAbs, Bispecific antibody; ADC, antibody-drug conjugate.

**Supplement Figure 3.**
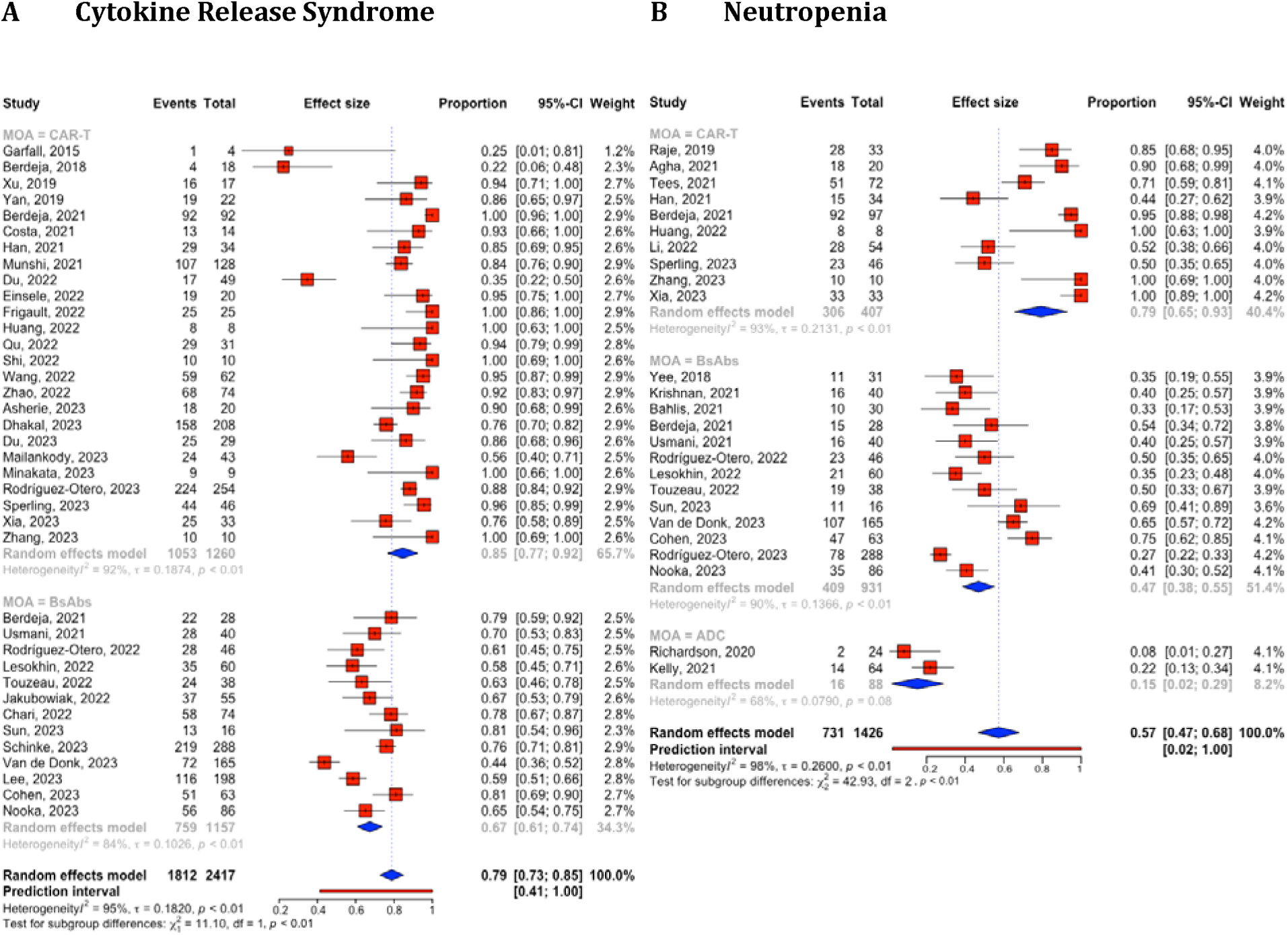
Combined quantitative analysis and subgroup analysis of cytokine release syndrome and neutropenia (≥Gr3) based on the therapies A. Cytokine Release Syndrome. B. Neutropenia (≥Gr3). CAR-T, chimeric antigen receptor T cell; BsAbs, Bispecific antibody; ADC, antibody-drug conjugate.

## Supplement Methods

### SEETrials

#### Pre-processing steps for abstracts collection and table conversion

We acquired raw abstract texts from each website. PubMed enabled direct saving in text format. Abstracts from ASCO and ASH websites were initially captured in a Word file and then converted to a text file (UTF-8) to facilitate the loading of a substantial number of abstracts for automated data extraction. Notably, annual conferences like ASCO and ASH permitted the inclusion of table-format data within abstracts. Acknowledging potential formatting challenges in documents containing tables, we developed an "Automatic Table Organizer" GPT model. This model effectively realigned columns and rows, addressing any formatting discrepancies. Specifically, for ASH conference abstracts, which presented tables in picture format, we employed Optical Character Recognition (OCR) to convert picture files into text files for further processing.

##### Automatic table organizer

A three-step process involved creating a text file with unorganized tables, running the "Automatic Table Organizer," and replacing the output with organized tables in original abstract documents.

##### Preparation of two abstract sets for one abstract with table

After running the Automatic Table Organizer, we observed enhanced extraction of table information but noted lower performance in extracting text information. To address this, we input two abstract sets into our GPT system: one with organized tables and the other without tables.

#### Knowledge ingestion

Prompts in specific areas often benefit from incorporating background knowledge within the prompt. We crafted tailored prompts that integrated oncology clinical trials’ knowledge to enhance the LLM’s analytical capabilities. For instance, we incorporated an understanding of primary and secondary endpoints for efficacy, and we provided safety outcome measurement with examples obtained through a thorough review of clinical trial protocols and manual abstract review.

The involvement of knowledge experts in this study encompassed various tasks: 1) identifying types of treatment efficacy and safety entities in cancer clinical trials, 2) formulating descriptions for the entire cohort, sub-cohorts, and each arm for different interventions or dosages specified in the prompts, 3) ensuring the clinical accuracy of the prompts, 4) providing feedback for prompt adjustments/calibration, and 5) manually reviewing system output to establish the gold standard for system validation.

#### Prompt modeling for data extraction

##### Prompt modeling

Using an initial prompt, we instructed our GPT system to extract and align information with cohort composition. Due to the token limit of the GPT-4 API model (8,192), we employed two separate GPT models for each of the comprehensive prompts: one for "Individual cohorts/arms and entire group" and " Individual cohorts/arms and subgroup group". Outputs from both models were combined in the subsequent Combining-processing step. Each prompt comprised three main components: a general instruction to extract study details, clinical outcomes, and adverse events; the abstract text; and a query. The query requested information on entities and values for specific tables, with detailed instructions for formatting.

##### Combining process with two GPT outcomes

To merge outputs from the "Individual cohorts/arms and entire group" and " Individual cohorts/arms and subgroup group" models, a prompt was created to accurately identify non-overlapping outputs, combine extracted information, and eliminate overlaps.

#### Postprocessing

We implemented a post-processing module to refine outputs from the data-extracting module by addressing two key formatting challenges: (1) addressing undesired allocations of entities and their corresponding values in the outputs and (2) ensuring that each cell within the value’s column contained a singular value, optimizing the presentation for computational analysis.

